# Impact of COVID-19 Outbreak on Healthcare Workers in a Tertiary Healthcare Center in India - A cross sectional study

**DOI:** 10.1101/2023.02.11.23285507

**Authors:** Shahzad Mirza, Arvinden VR, Mercy Rophina, Jitendra Bhawalkar, Bhavin Chothani, Uzair Khan, Shivankur Singh, Tanya Sharma, Aryan Dwivedi, Ellora Pandey, Shivam Garg, Mukhida Sahjid Sadrudin, Zeeshan Shabbir Ahmed Sange, Shalini Bhaumik, Jessin Varughese, Vishwamohini Yallappa Devkar, Jyoti Singh, Anju mol V K, Veena K, Husen Shabbir Husen Mandviwala, Vinod Scaria, Aayush Gupta

**Author notes:** Address for correspondence C-2002, Empire Square Society, Chinchwad, *, **Corresponding author Email:**. Shahzad Mirza is an Associate Professor in the Department of Microbiology and Hospitals Infection control officer. His research interests include antibiotic resistance, antimicrobial stewardship, hospital-associated infections, and clinical Microbiology.

## Abstract

Numerous speculations have continually emerged, trying to explore the association between COVID-19 infection and a varied range of demographic and clinical factors. Frontline healthcare workers have been at the forefront of this illness exposure. However, there is a paucity of large cohort-based association studies, performed among Indian health care professionals, exploring their potential risk and predisposing factors. This study aims to systematically utilize the demographic and clinical data of over 3000 healthcare workers from a tertiary hospital in India to gain significant insights on the associations between disease prevalence, severity, and post-infection symptoms.

**Article Summary:** Potential associations between various demographic and clinical factors with disease severity and post COVID syndromes among a large cohort of healthcare workers in India suggest that Smokers were discovered to be more vulnerable to COVID-19 infection because of their immunocompromised lung health and Blood group B, like previous studies, was found to possess an increased risk of predisposition to long COVIDs

## Introduction

The index case of COVID-19 was reported on January 30, 2020, in India ^1^. This was followed by a sharp rise in the number of cases, leading subsequently to the adaptation of various combat strategies based on the population as well as the healthcare infrastructure of the nation ^2^. Although most COVID-19 patients had mild to moderate symptoms, some patients, particularly those with underlying comorbid conditions and/or elevated levels of inflammatory markers, went on to develop serious disease and/or passed away (Mammen et al. 2021). Gradually, a handful of studies started reporting the presence of prolonged COVID-19 symptoms like fatigue, cough, shortness of breath, insomnia, mood disturbances, anxiety, and myalgias in a variable proportion of patients ^3^,^4^. The World Health Organization (WHO) estimated about 10-20% of infected patients to suffer from “long covid” ^5^

Since its onset, India has reported over 43.2 million cases of COVID-19, making it the second most affected country in the world (following the USA). As of June 2021, the cumulative COVID 19 deaths in India were estimated to be more than 4 million ^6^.

Healthcare workers (HCW), due to their frontline nature, are at the greatest risk of contracting or spreading COVID-19 infection. The prevalence of infection among HCWs has exceeded 10% in Italy ^7^, and a total of 9282 HCWs were confirmed with COVID-19 as of April 9, 2020, in the United States ^8^. In India, the ICMR portal instituted to capture information of individuals undergoing testing for SARS-CoV-2 infection found 5% of HCWs to be positive for COVID-19 after an analysis of 21,402 records ^9^. Though the association of various clinical characteristics with the outcomes of COVID-19 has been previously reported in the Indian population, a representative study among HCWs is substantially lacking. This study serves as the first of its kind to perform a large-scale statistical analysis on a range of clinical and demographic characteristics of over 3000 HCWs in a tertiary care Indian hospital and reports rates of vaccinations, breakthrough infections, and reinfections along with significant predictors of long Covid.

## Materials and methods

### Study population and design

This cross-sectional study was carried out over a period of 12 weeks (July - October, 2021) after obtaining institutional ethical clearance and informed consent - on all consultants, postgraduate and undergraduate students, interns, nurses, nursing auxiliaries, clerical staff, security personnel, administrative staff, pharmacists, and cleaners working in a tertiary care hospital that served as a dedicated 850 bed covid care hospital in western India. A 43-item survey was designed using Google Forms and piloted to assess the design as well as check the feasibility and validity of the questions. A team of postgraduate, undergraduate students and nurses conducted face to face interviews and filled in information in the form. Vaccination certificates as well as SARS-CoV-2 RT-PCR reports were manually verified. Post COVID-19 symptoms, also known as “long COVID,” was defined as persistent symptoms 3 months from the onset of COVID-19 with symptoms that lasted for at least 2 months and could not be explained by an alternative diagnosis ^10^ were also duly included.

### Dataset used in the study

The study dataset comprised demographic and clinical information of over 3000 healthcare workers, including age, gender, height, weight, blood group, vaccination status, severity of infection, history of hospitalization, long covid symptoms and lifestyle habits like smoking, use of tobacco, alcohol, diet, and other comorbidities if any.

### Estimation of statistical association

Univariate Fisher exact tests were performed to ascertain the association of long covid with various parameters like age, sex, comorbidities, vaccination, reinfection, BMI index, and blood group. Multiple test corrections of p values were performed using the Bonferroni method. A multinomial regression model was generated for analysis of the severity of COVID-19 and its association with all the above-mentioned parameters using the multinom function from nnet (version 7.13-16) ^11^ R package. p values were calculated from t statistics and multiple test correction was performed. All data analysis and visualization was performed in R (version 4.1.2) ^12^ using the ggplot2 package (version 2.3.3.5)^13^.

## Results

### Data formatting and analysis

#### General demographic data

The study enrolled a total of 3329 HCWs belonging to over 20 departments. The mean age of the study group was 28.11 (± 8.95) years, with 2110 (63.38%) females and 1219 (36.62%) males. Undergraduate students (n = 849, 25.50%) and nurses (n = 821, 24.66%) comprised the major proportion of the dataset. Chefs and clerks were included under the administrative category and infection control staff in the nurse’s category. Majority of the participants had no history of smoking, tobacco chewing or alcohol drinking. Blood groups B (1095, 32.89%) and Rh positive (3003, 90.21%) were found to be the most common. 75.3% of the total enrolled were found to possess no clinical comorbidities, whereas asthma (57, 1.71%), diabetes (54, 1.62%), hypertension (64, 1.92%), stroke (1, 0.03%), hypo- and hyperthyroidism (68, 2.04%) were reported by the rest.

#### Vaccination status

78.6% (2618) of the study population were found to be vaccinated at least once with either Covishield (2295), Covaxin (303), Pfizer (7), Covovax (6), Sinopharm (5) or Sputnik (1), while the rest (711, 21.4%) were unvaccinated. There were multiple reasons reported for not taking a vaccine, including pregnancy, allergies, unavailability of slots and personal disinterest. Undergraduate students (91.76%), residents (92.61%), interns (96.64%) and faculty (91.19%) were found to be vaccinated while nurses (68.90%), technicians (63.48%) and other admin staff (69.13%) were vaccinated in lower percentages as compared with the prior groups as depicted in **Figure 1**. No serious adverse effects following immunization (AEFIs) were observed in the current study.

**Figure 1.**
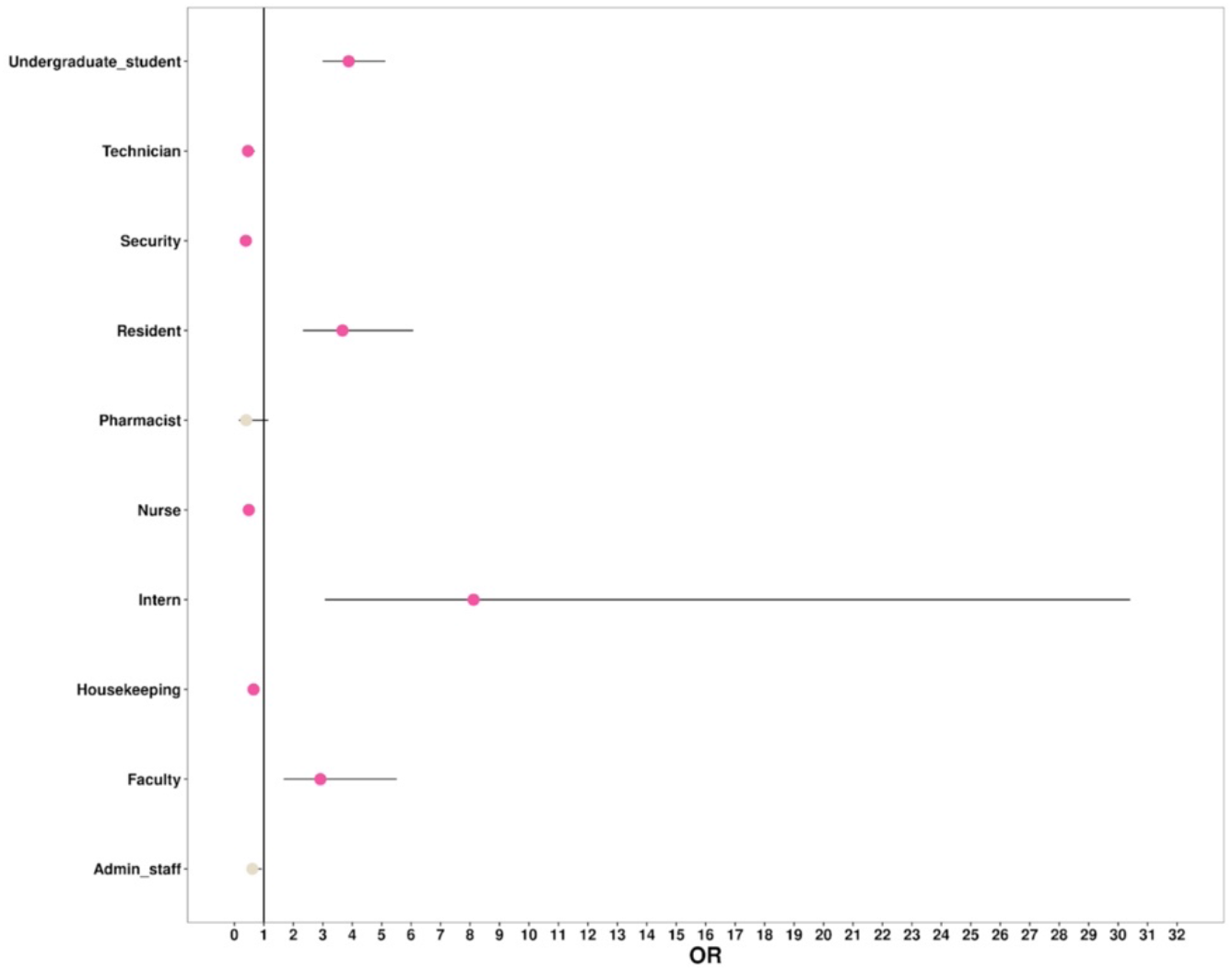
Schematic representation of the statistical association of vaccination status and occupational categories of healthcare workers enrolled in the study.

#### Status of infection and the severity of outcomes

654 (19.65%) HCWs were found to be SARS-CoV-2 positive at least once, of which a small proportion (n=98,15%) did not get their infection confirmed by RT-PCR or antigen tests despite being symptomatic. **Figure 2**. provides a schematic representation of the status of infection among various occupational categories of health care workers enrolled in the study. Fever, cough, body ache, breathing difficulties, loss of smell and taste (symptoms typically observed during the delta wave) were the most reported symptoms. 75.1% (n=491) of the infected HCWs did not require hospitalization, whereas the rest (24.6%, n=161) were hospitalized for an average duration of 9 days. Oral and injectable antibiotics were the most administered treatments, followed by favipiravir, steroids, anticoagulants, supplemental oxygen, and remdesivir, as shown in **Figure 3**. Based on symptoms and the history of hospitalization, infected patients were categorized as severe (needing ICU admission or intubation), moderate (general hospitalization) and mild (symptomatic but under home isolation). Of the total participants enrolled in the study, 55 (8.41%) were found to be asymptomatic, while 571 (87.31%), 23 (3.52%), and 5 (0.76%) experienced mild, moderate, and severe COVID-19 respectively. Although hypothyroidism and reinfection were found to be significantly associated with moderate and severe infections, owing to their minimal counts, the exact reliability of these results could not be ascertained.

**Figure 2.**
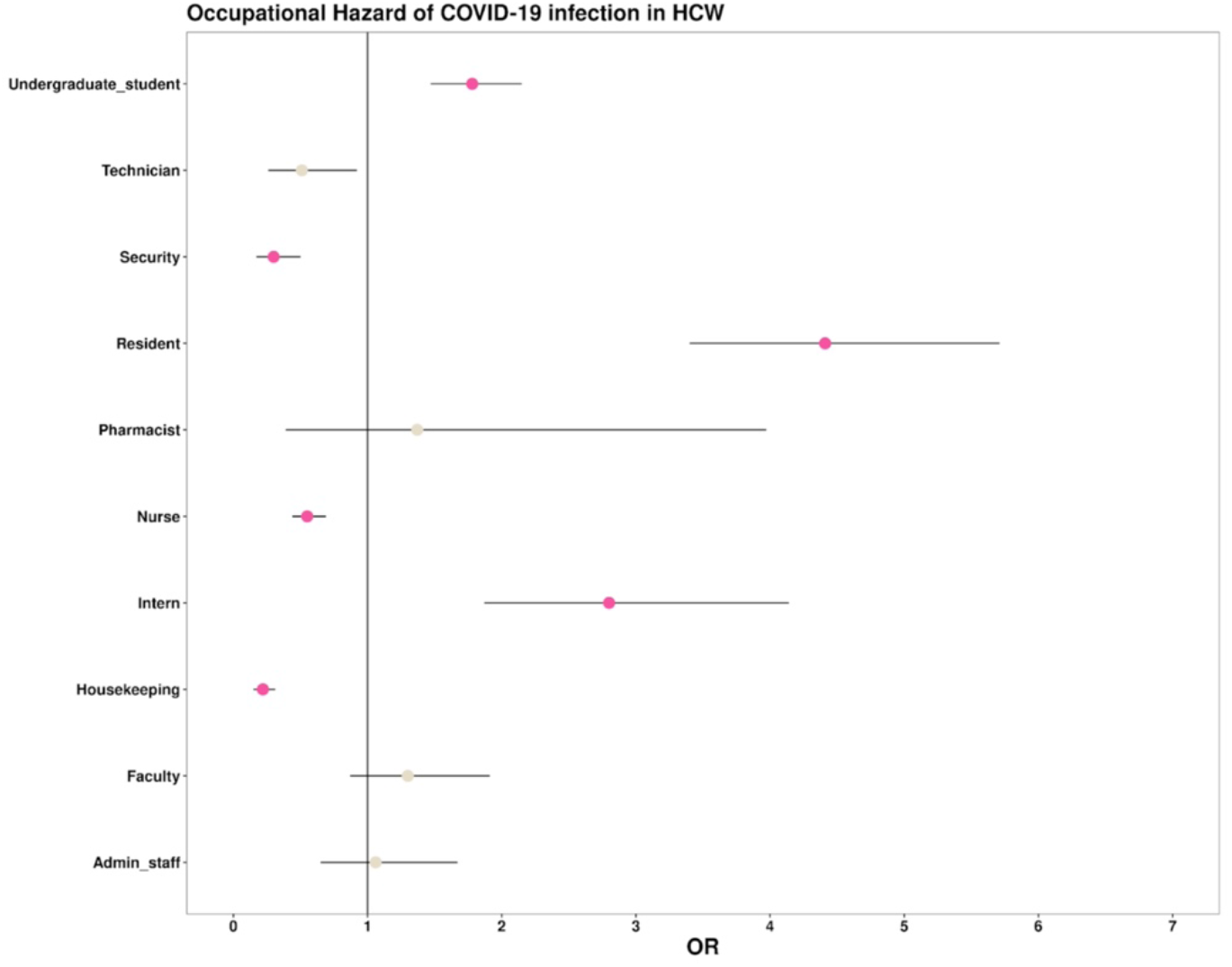
Schematic representation of the statistical association of status of infection and occupational categories of healthcare workers enrolled in the study.

**Figure 3.**
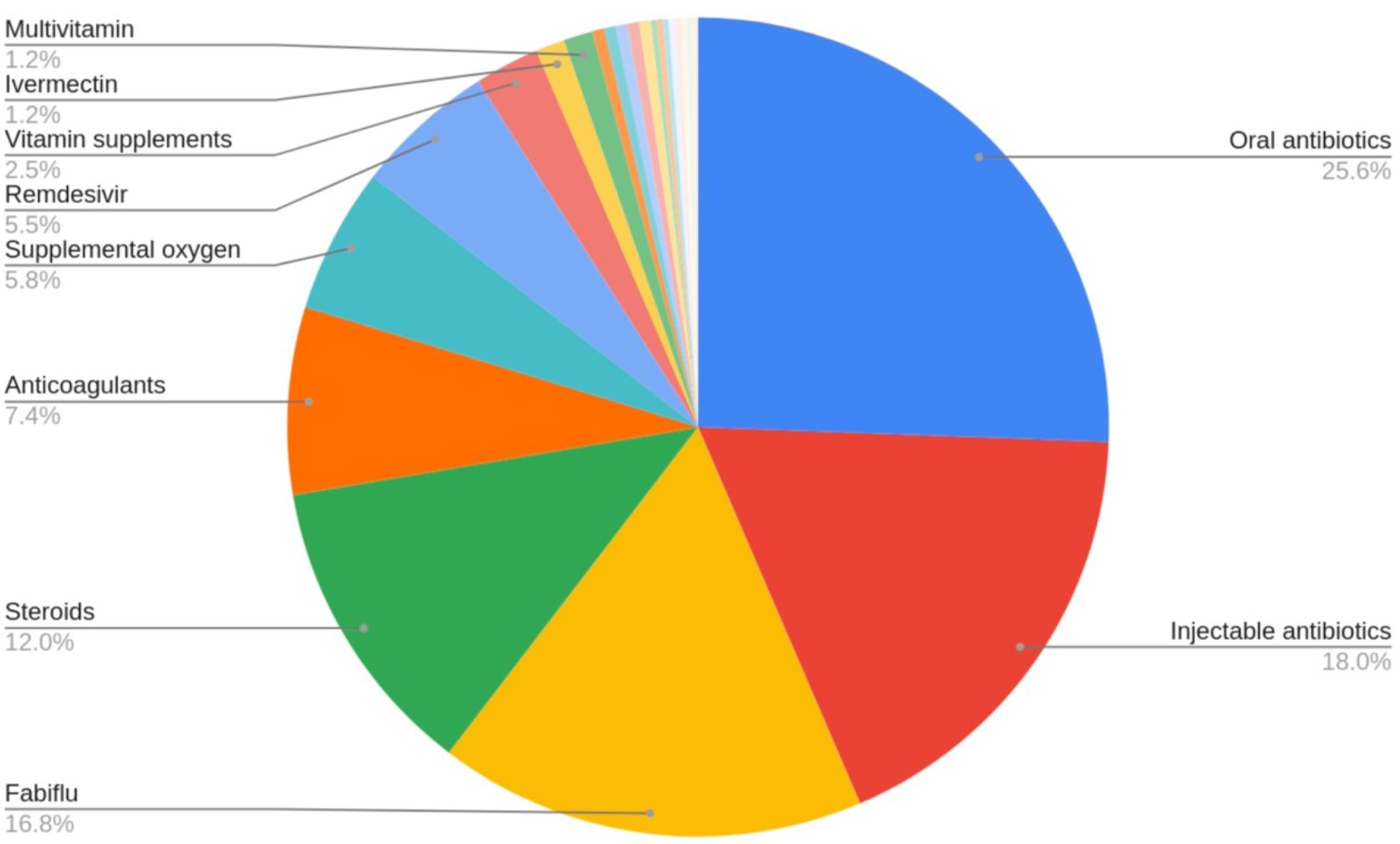
Schematic representation of the proportion administration of various COVID-19 treatments.

A total of 165 (25.23%) cases had breakthrough infections of which 147 (single dose: 38, double dose: 109) took Covishield and 18 (single dose: 8, double dose: 10) took Covaxin. Symptoms due to breakthrough infections were largely mild with 16 and 145 individuals demonstrating asymptomatic or mild infections, while two each had moderate and severe disease. The mean time for breakthrough infection from the time of the first vaccine dose was found to be 61.89 days (range: 2 to 281 days). No significant difference was found between partially or fully vaccinated when breakthrough infection time was compared from the day of initial vaccination.

In addition, 25 cases of reinfection were also observed in our study population. Surprisingly, all reinfections were found in vaccinated individuals (6 (24%), partially vaccinated; and 19 (76%), fully vaccinated). Further, 14 individuals with reinfection (56%) also reported the presence of long COVID symptoms.

### Overview of long covid analysis

The survey had listed a range of long covid symptoms along with an open-ended question to gauge the variability of symptoms. 206 (6.19%) individuals were found to be suffering from long COVID. Persistent weakness/tiredness, lasting from 12 weeks to 6 months, was the most experienced post COVID symptom. Asthenia, loss of smell, myalgia, headache, neurotic symptoms, shortness of breath, loss of appetite, menstrual abnormalities, cough, joint pain, sore throat, frequent sleeping troubles, difficulties in concentration/confusions and leg pain were found to be the other common long covid symptoms. A handful of cases with rectal bleeding, weakness in the eyesight, and panic attacks were also seen. The distribution of these symptoms is shown in **Figure 4**.

**Figure 4.**
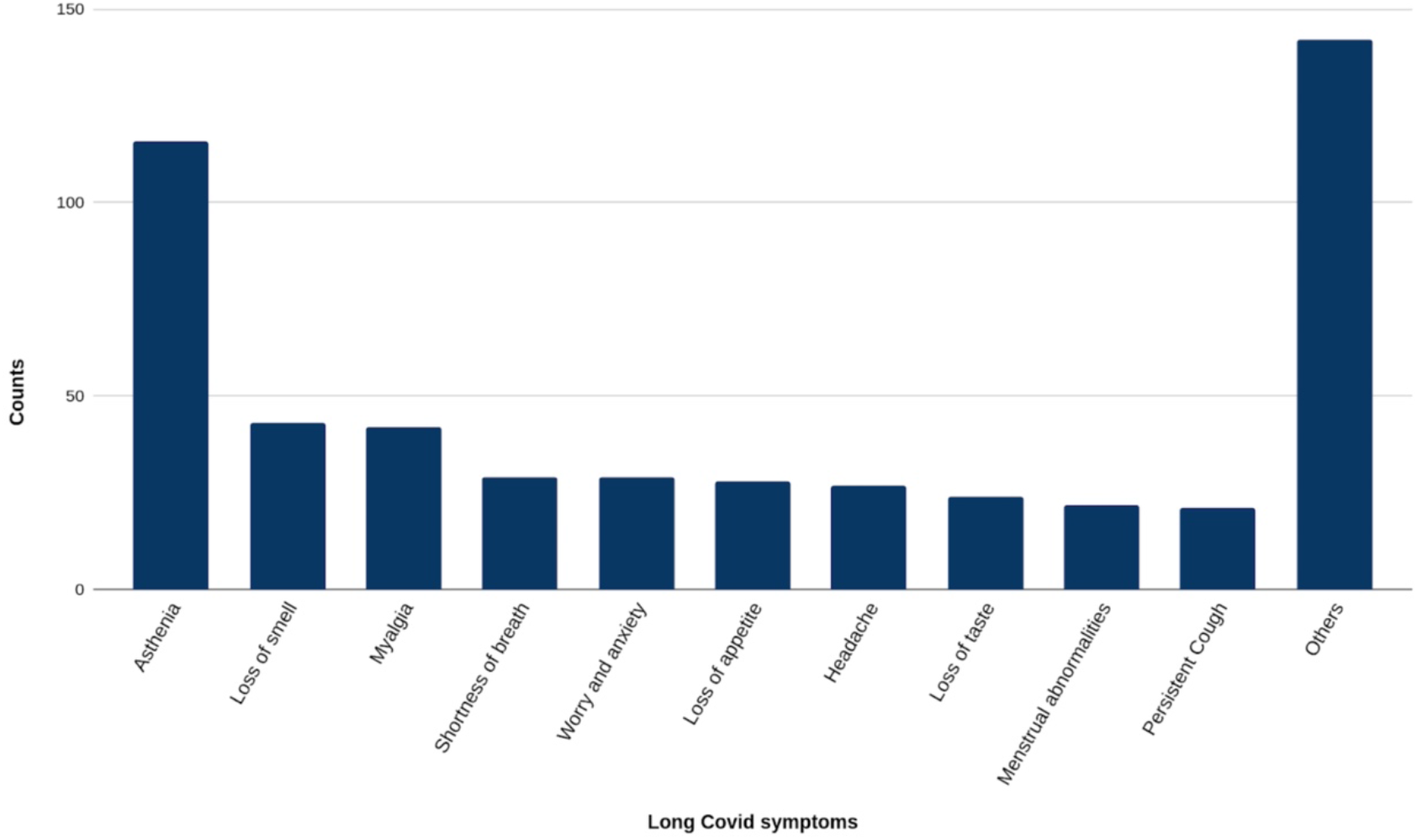
Distribution of various long covid symptoms observed in the study.

Surprisingly, undergraduate students (50%) who were attending classes remotely for a period of time were found to experience long covid symptoms more commonly, followed by nurses (14.08%) and residents (11.65%) as compared to the rest of the study group. The precise summary of the prevalence of long covid among various age groups and occupations is schematically shown in **Figure 5**.

**Figure 5.**
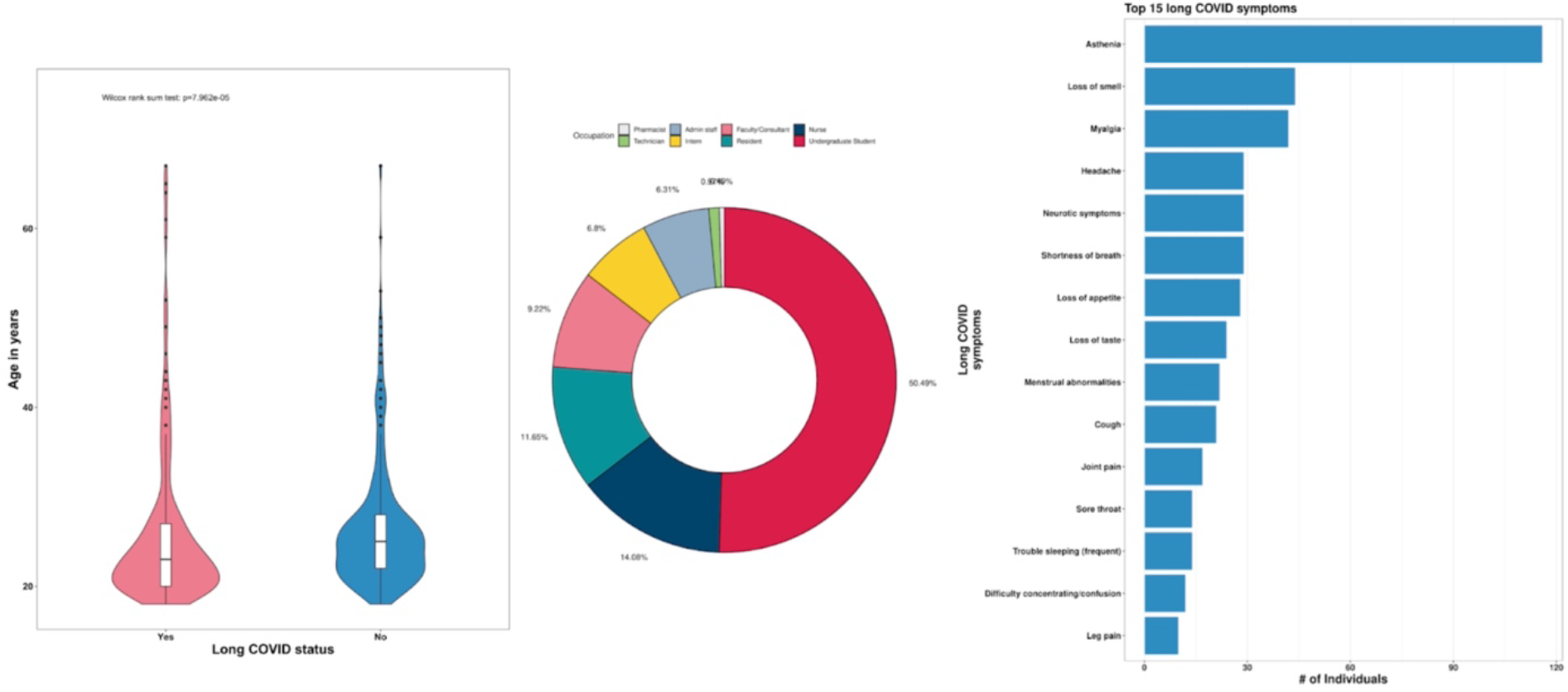
Schematic representation of the status of long covid among various age groups and populations along with the highly prevalent long covid symptoms.

We then tried to identify the presence of significant associations between long covid and various comorbid conditions, lifestyle factors, demographic factors, blood group, vaccination status, and history of infections. On univariate analysis, it was found that females possessed a significantly higher risk of long covid in comparison to males. In addition, drinking alcohol (22, 10.7%) and blood group B (65, 31.1%) were identified as significant risk factors for long covid in healthcare workers. No significant association was observed between the severity of the infection and the onset of long COVID as shown in **Figure 6**. A summarized tabulation of the demographic and clinical details is provided in **Table 1, while Figure 7** summarizes the significant potential predictors and risk factors of long covid. Table 2 provides a complete list of observed P-values and Odds ratios for various associations performed in the study

**Table 1.**
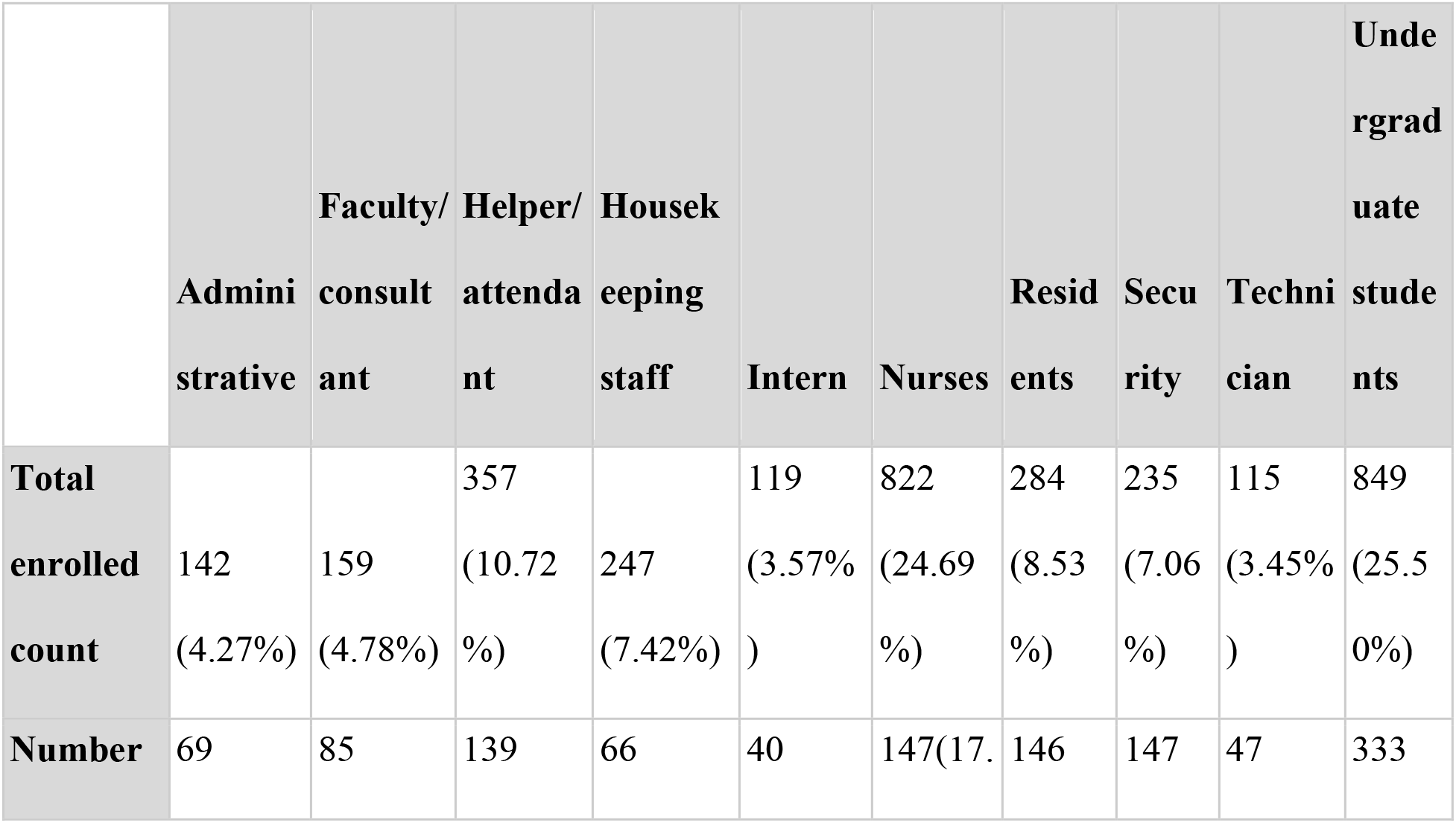
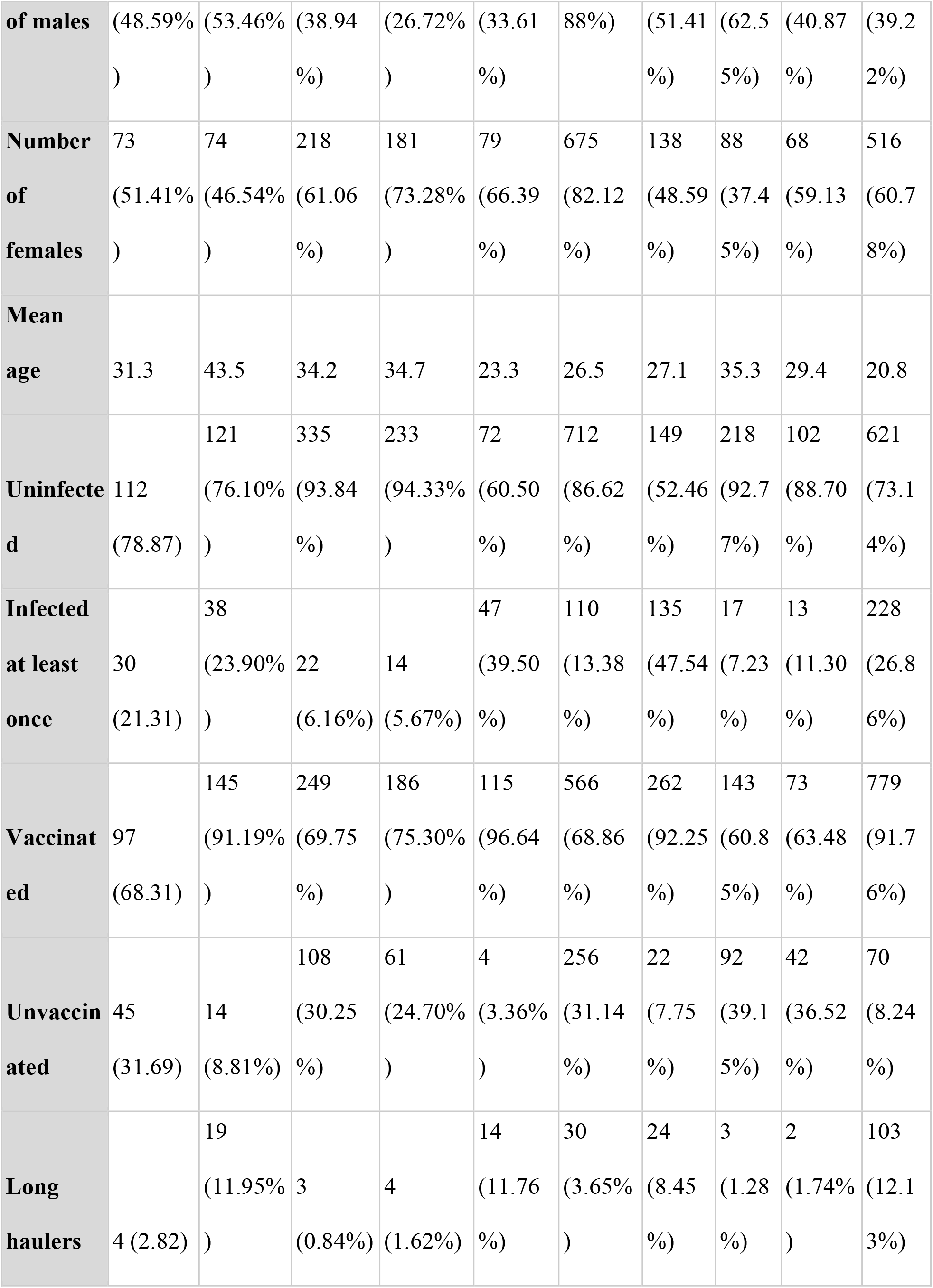
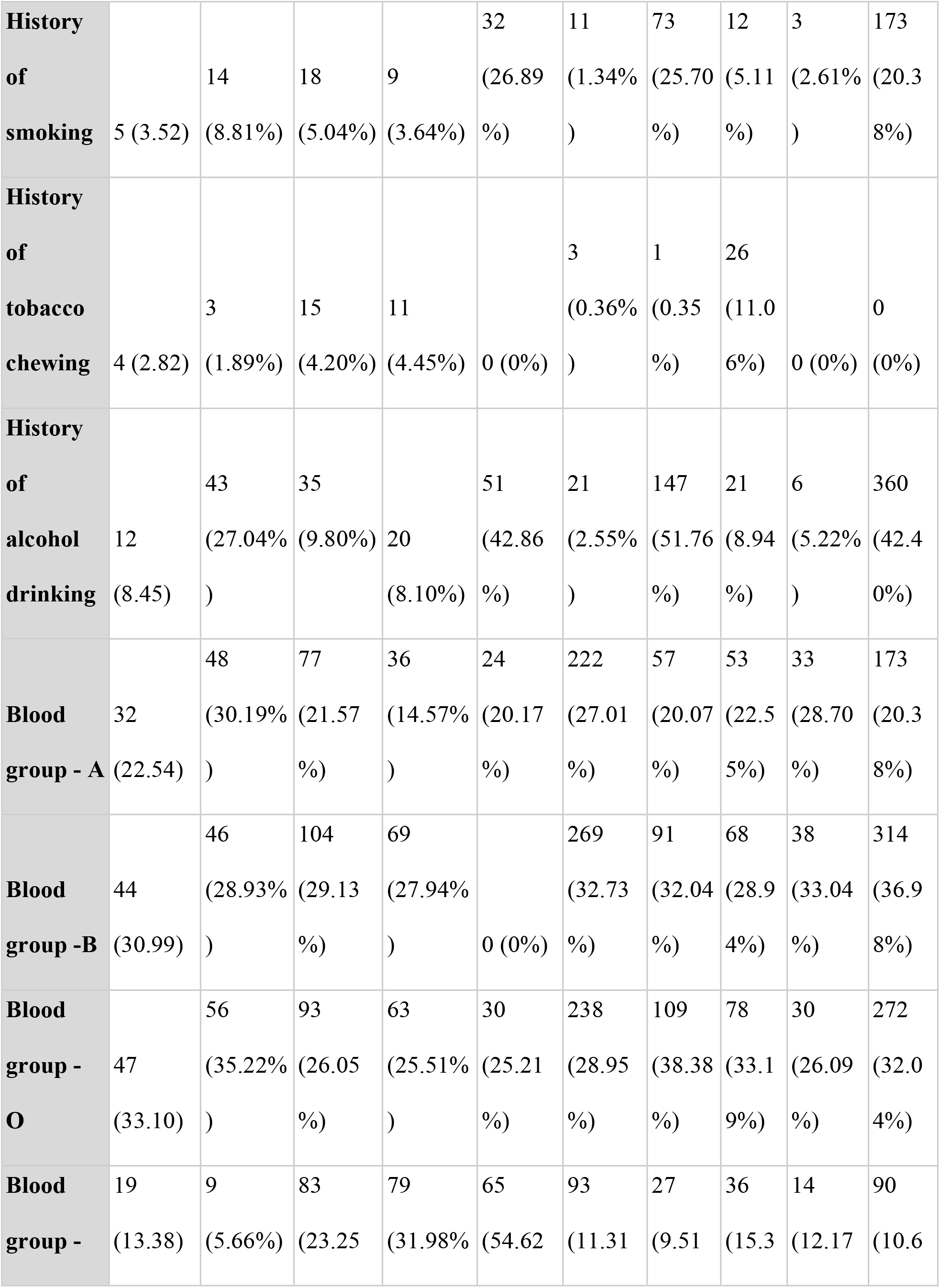
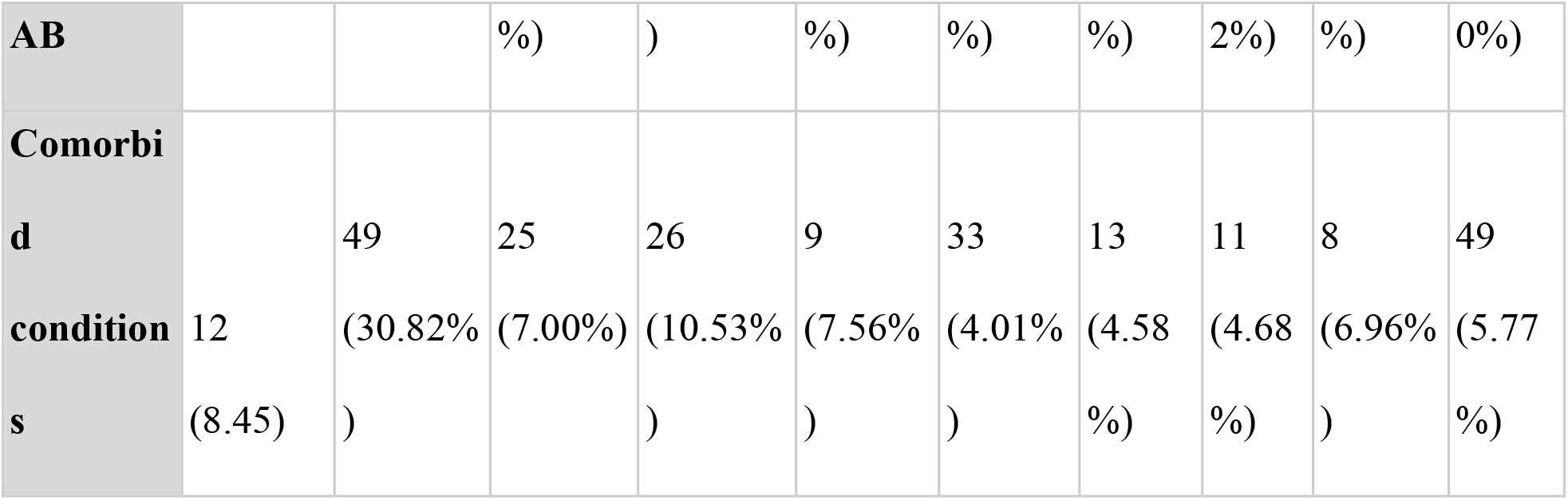
Tabulation of clinical and demographic details of study datasets.

**Table 2.**
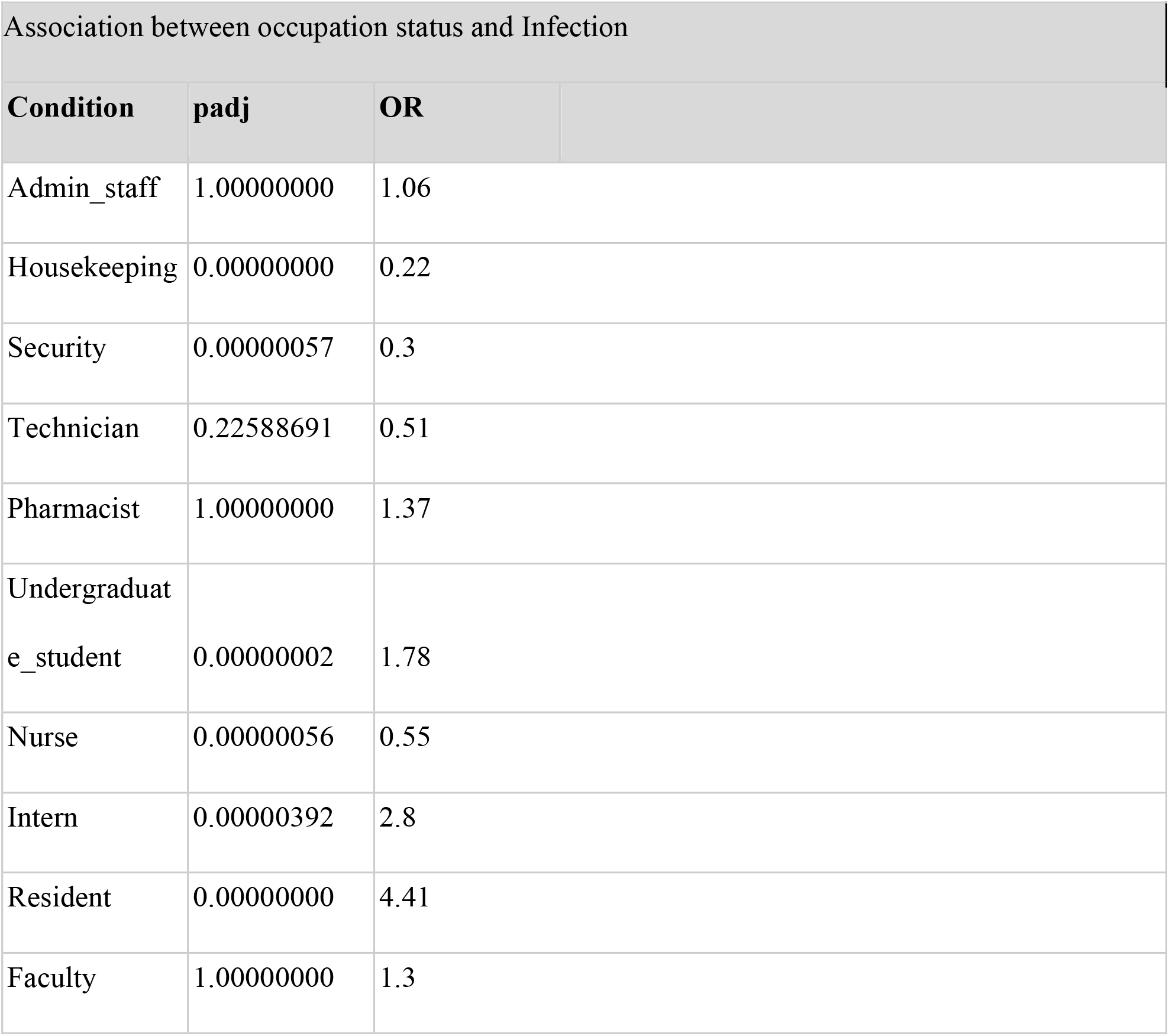
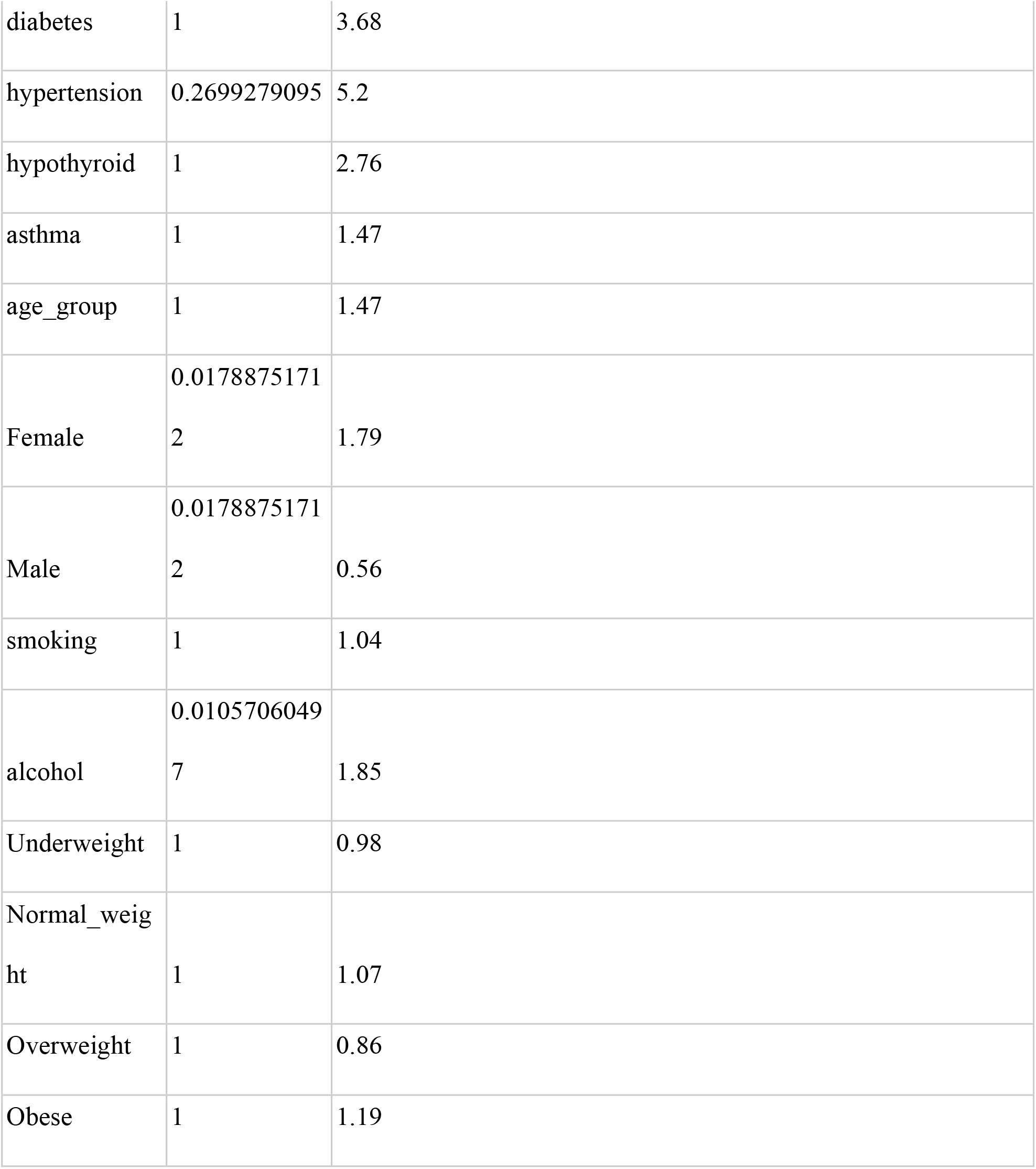
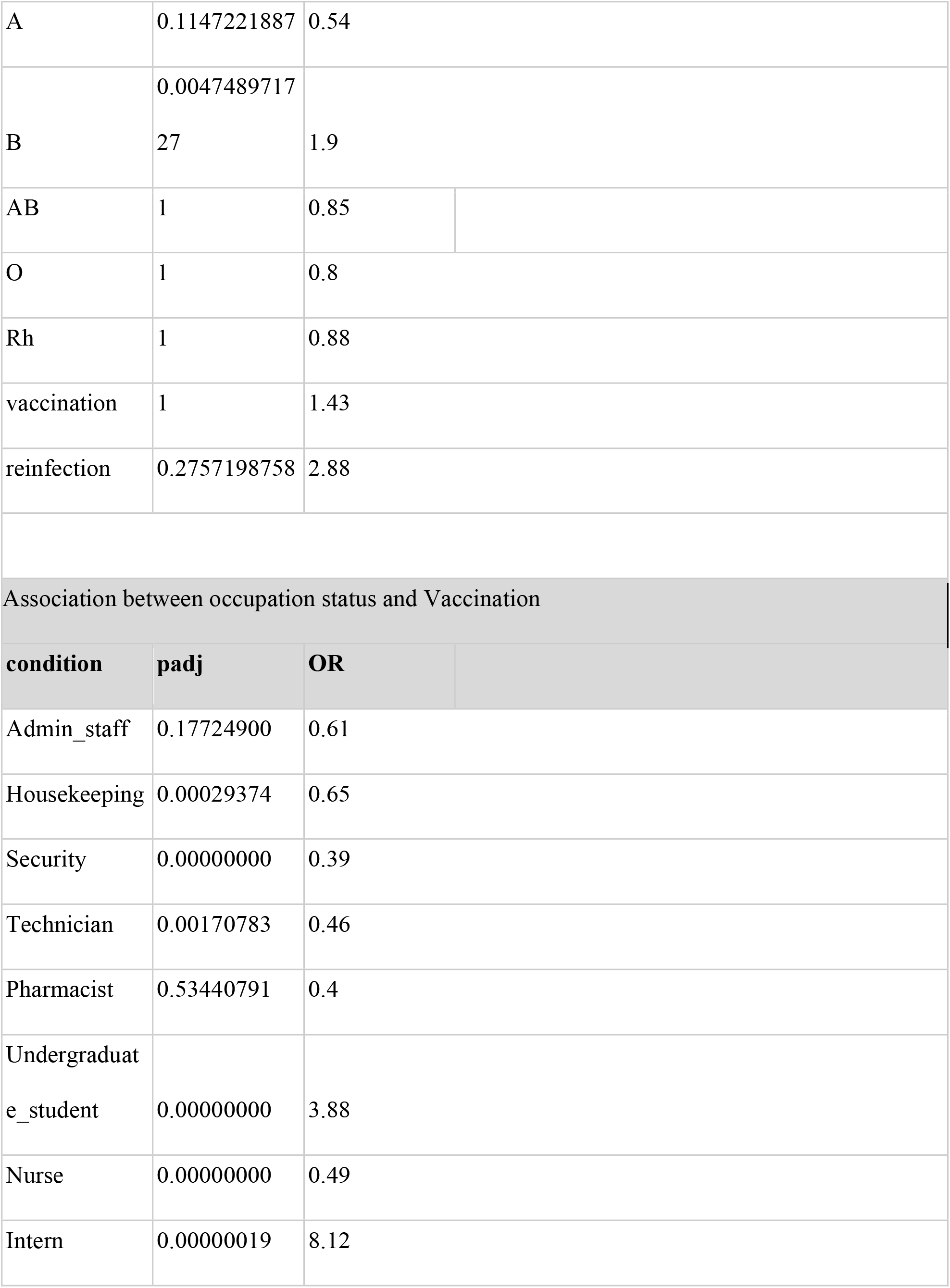
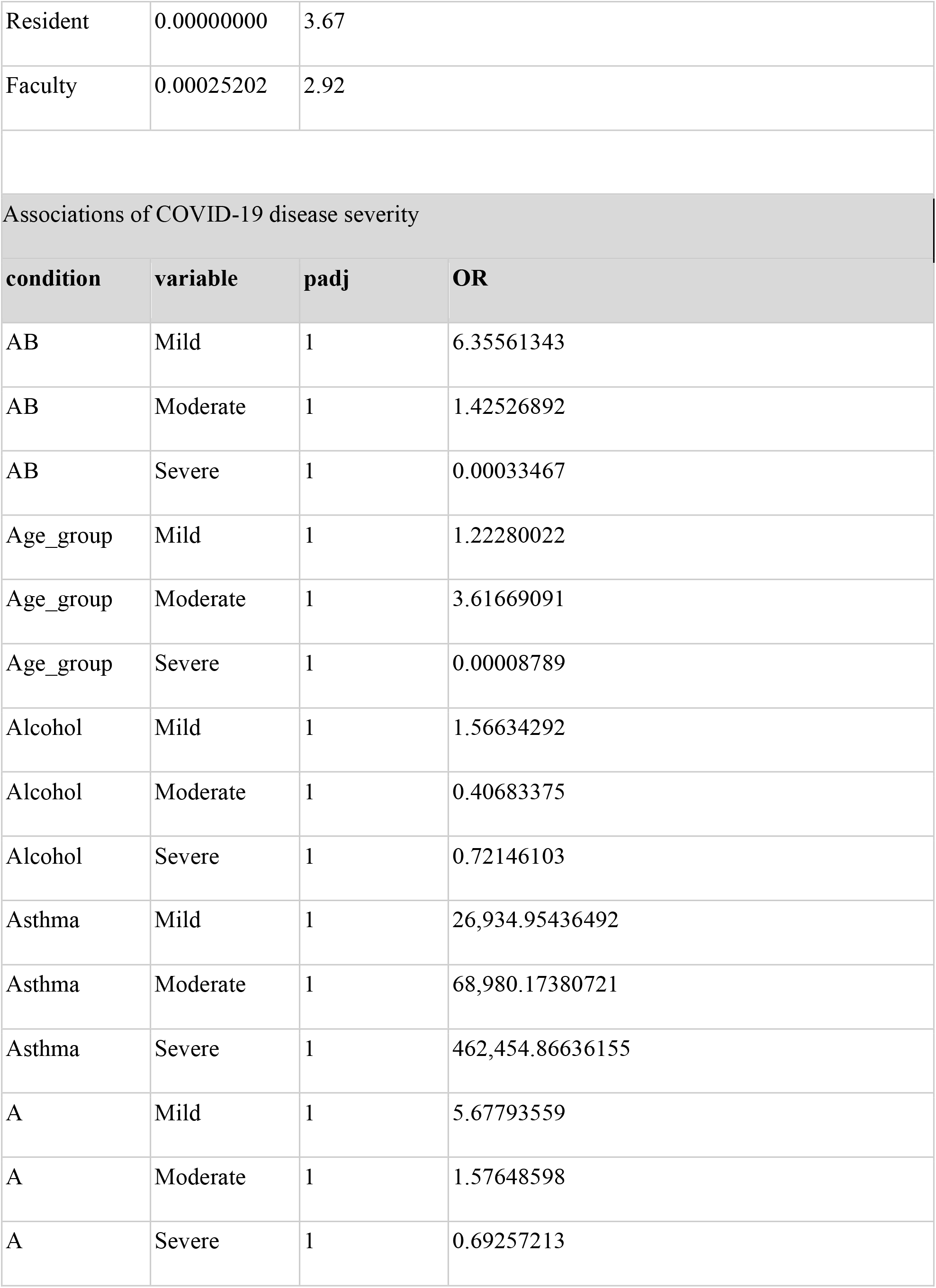
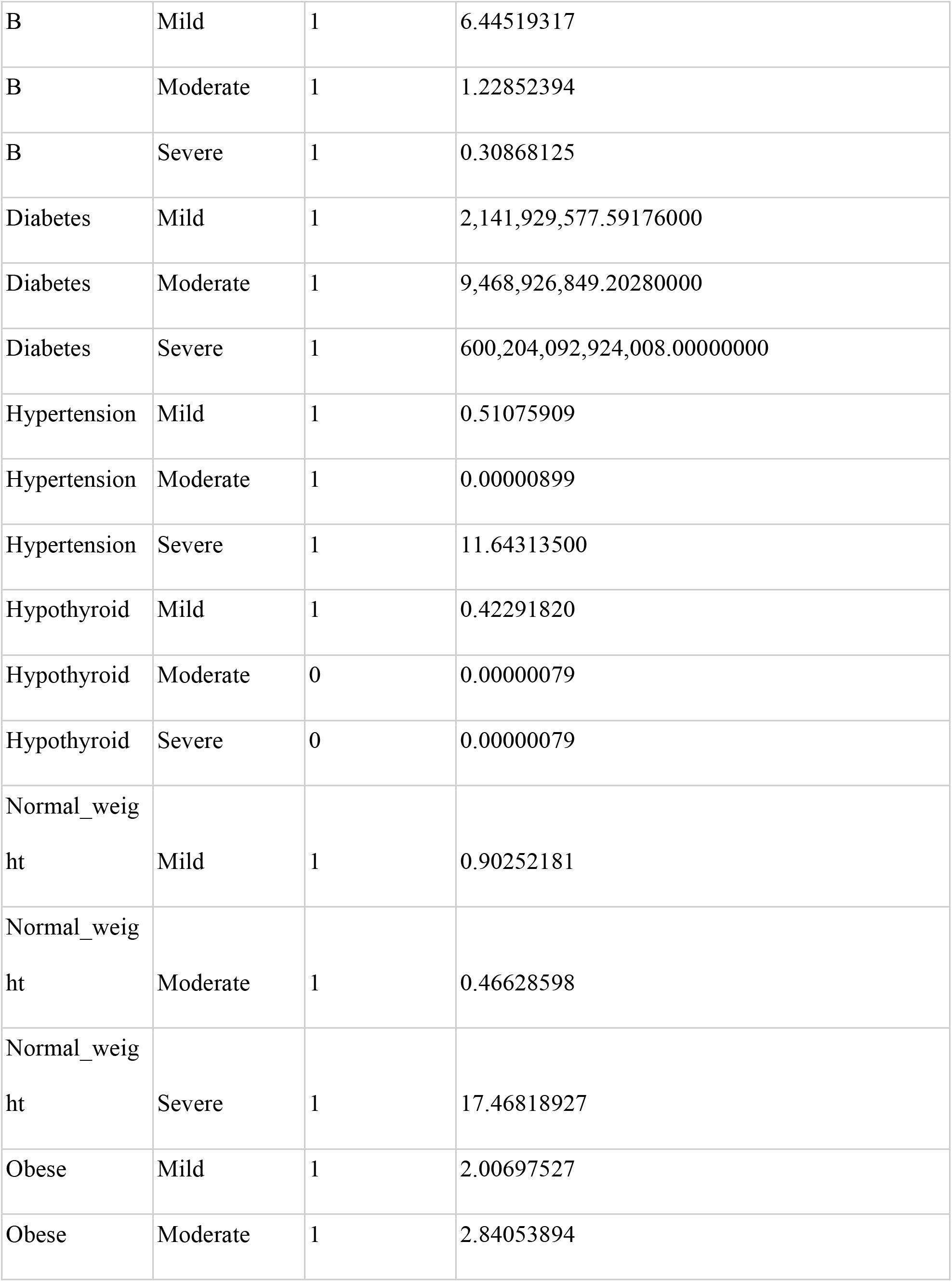
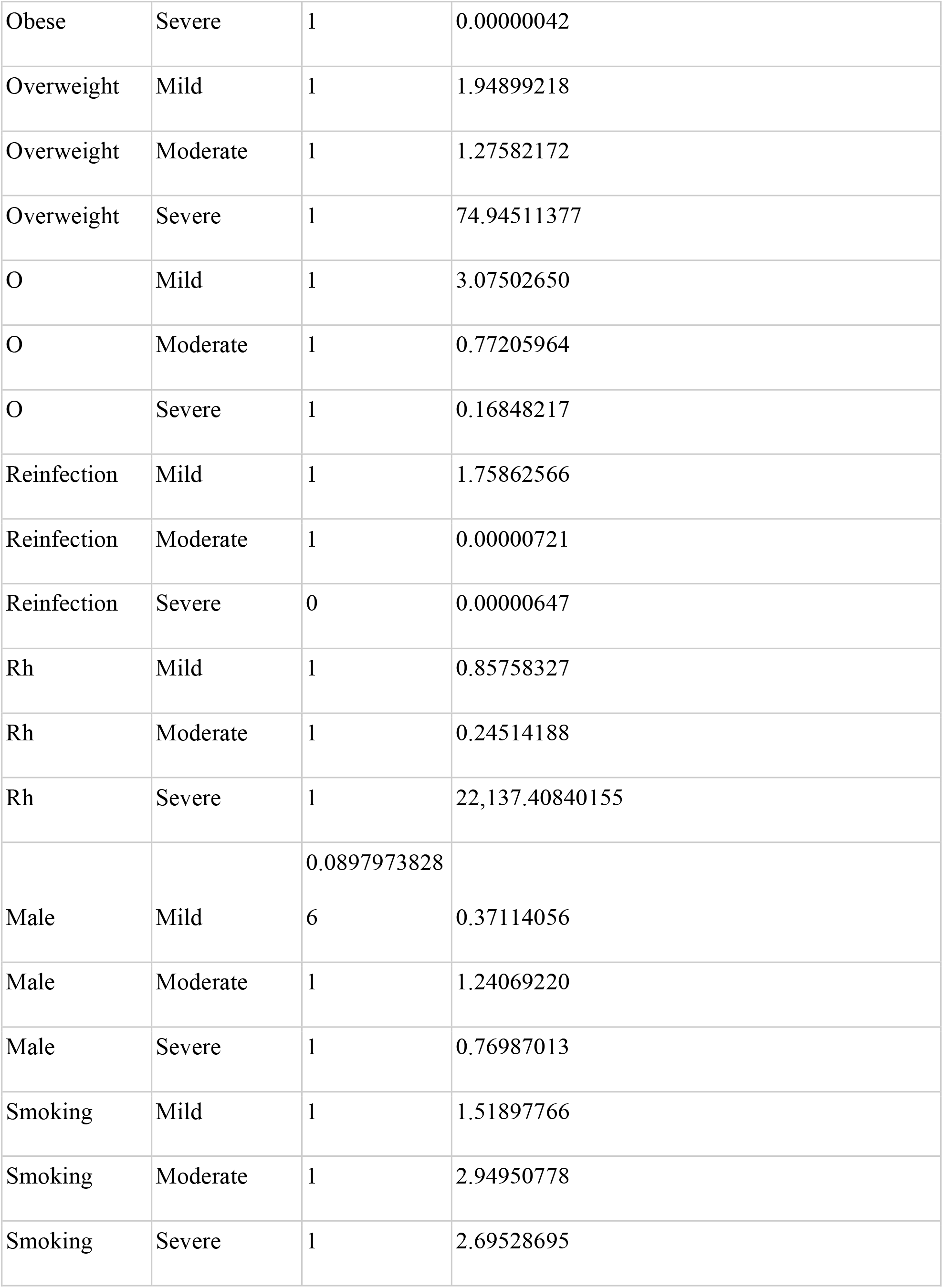
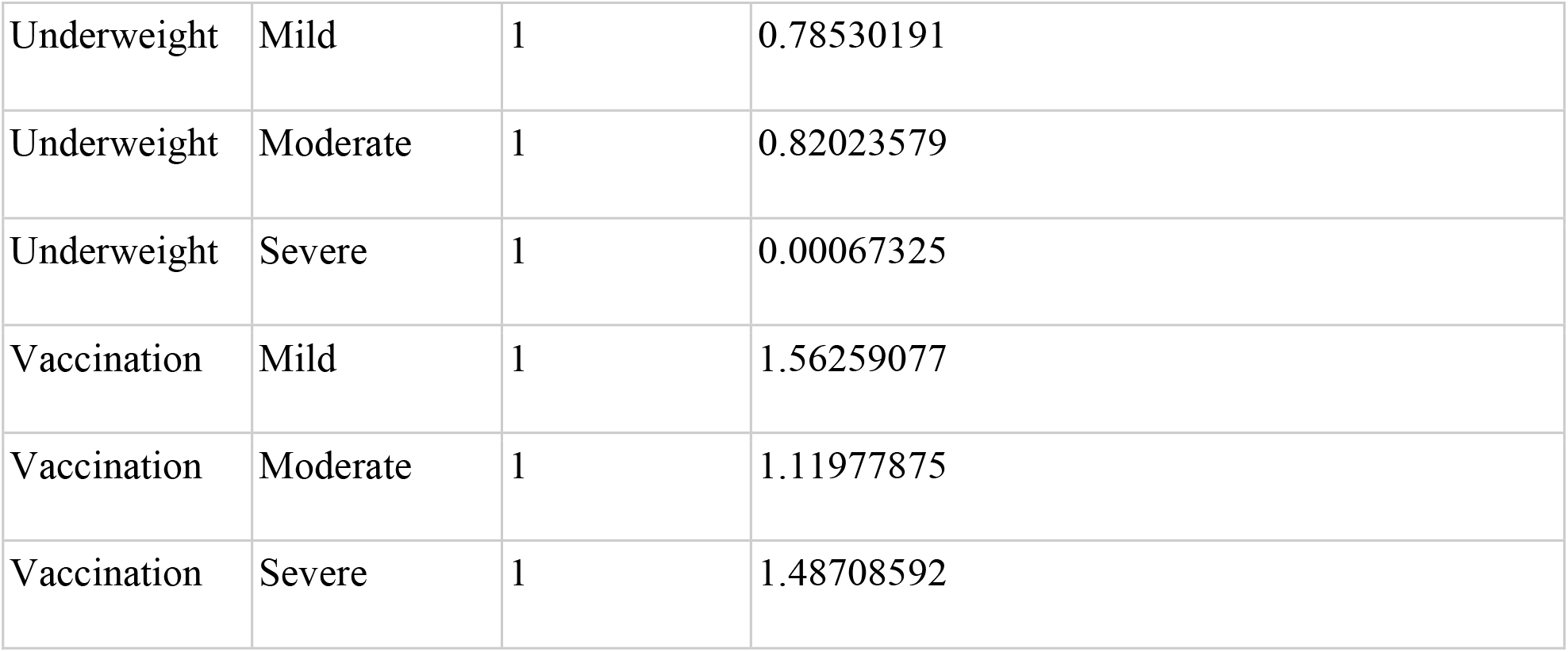
The observed P-values and Odds ratios for various associations performed in the study.

**Figure 6.**
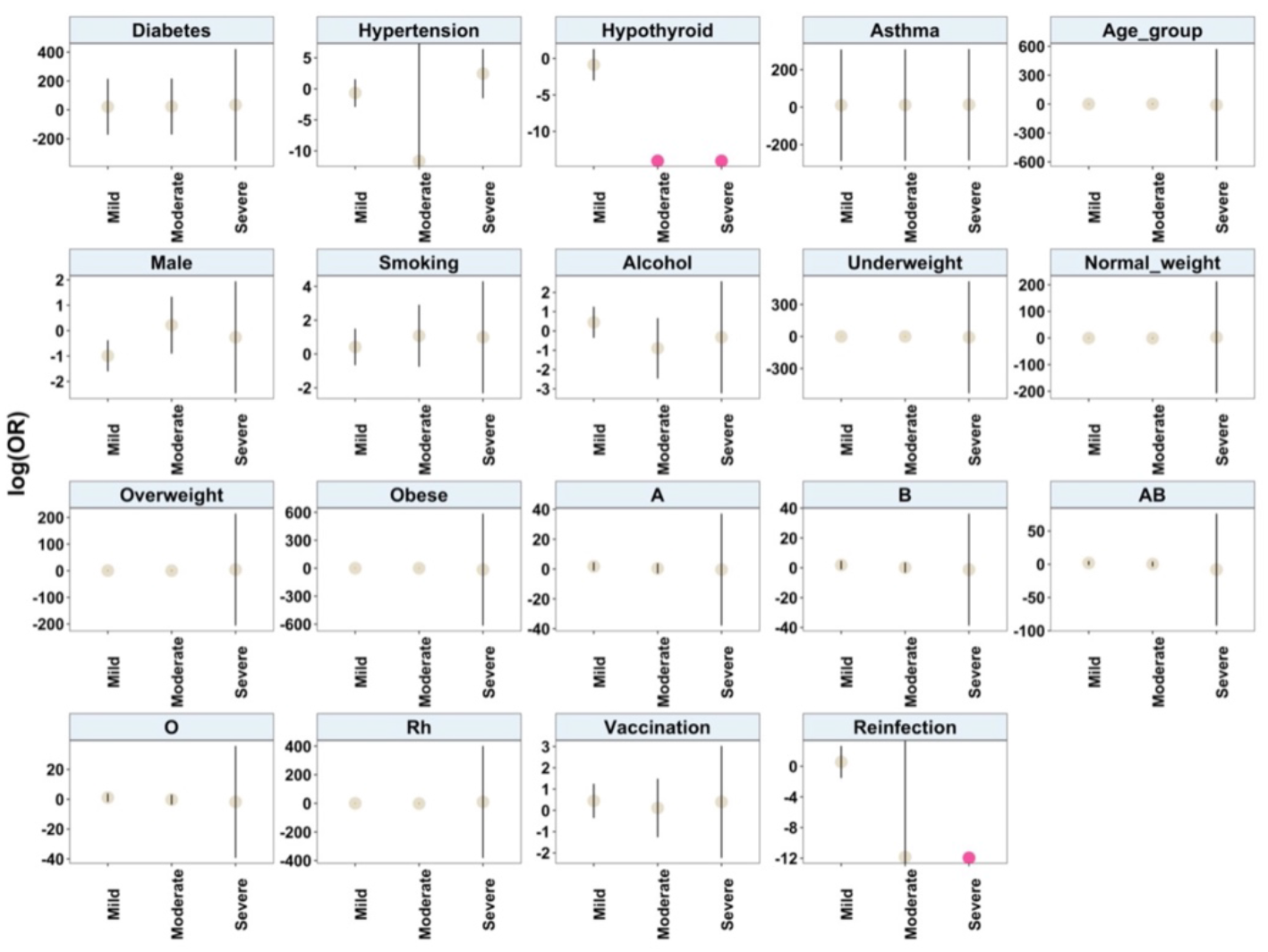
Statistically significant potential predictors and risk factors of moderate and severe COVID-19 outcomes.

**Figure 7.**
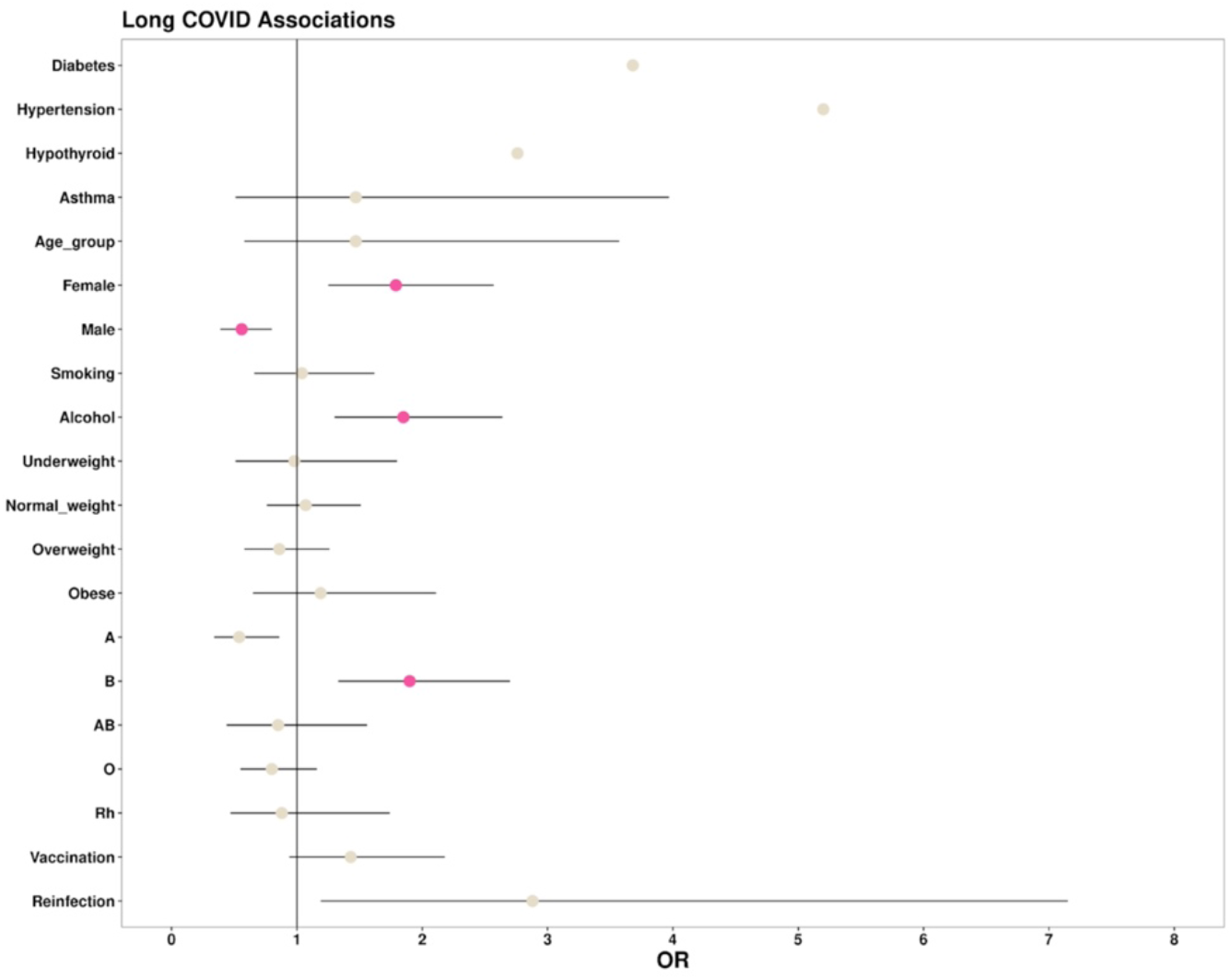
Statistically significant potential predictors and risk factors of long covid.

#### Temporal trends of COVID-19 risk in healthcare workers

With the aim of identifying significant differences in the susceptibility to COVID-19 infections between the general population and healthcare professionals, a temporal trend analysis was performed to compare the infection rates reported in Pune from February 2020 to October 2021 with the infection rates estimated from the study dataset. Information on the daily reported COVID-19 cases from Pune was fetched from <u>Covid19tracker.in</u>. Our analysis revealed that there were no significant differences in the trend of COVID-19 onset between the general population and healthcare workers. An overview of the similarity observed in the infection rates is shown in **Figure 8**.

**Figure 8.**
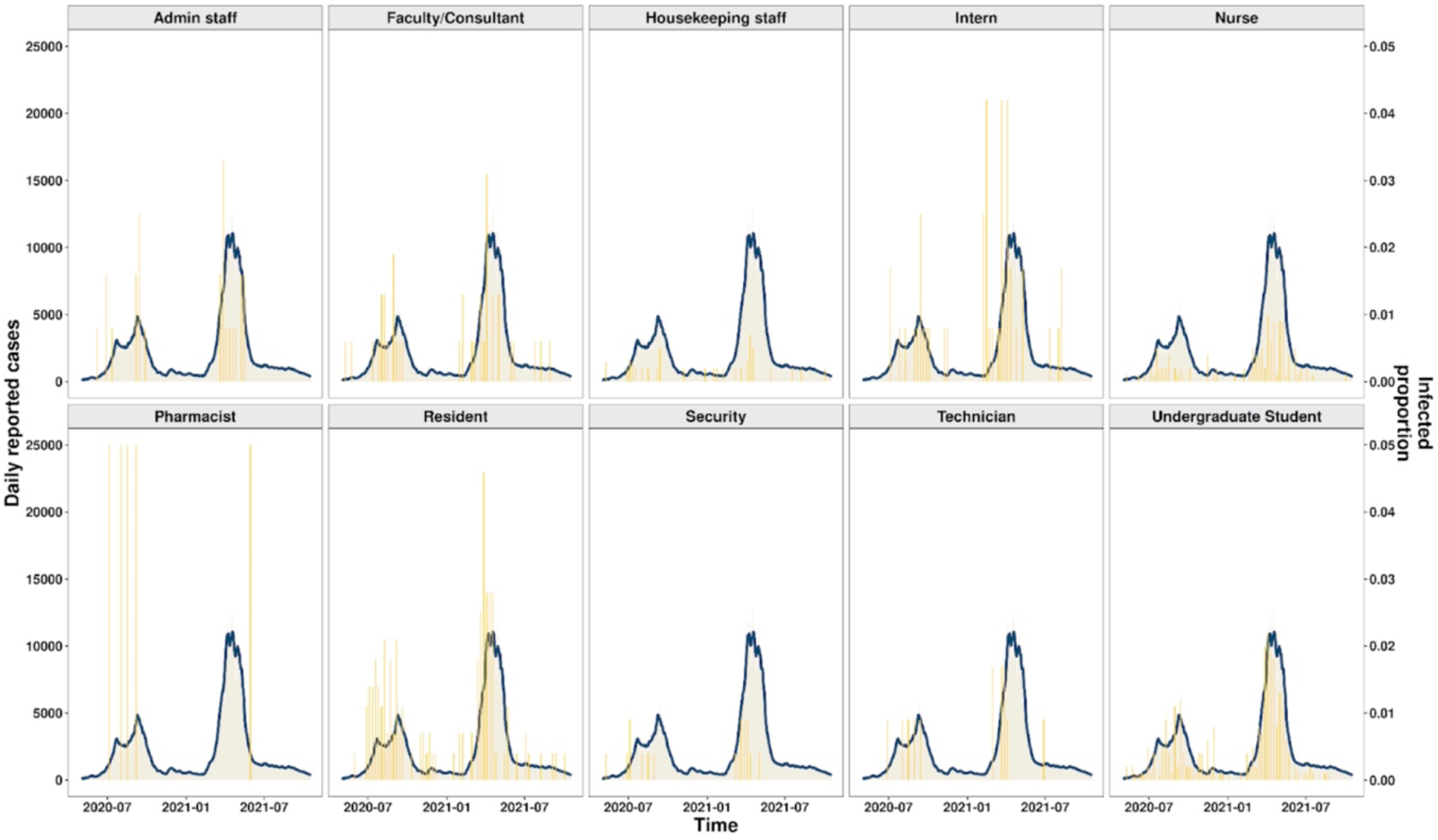
Overview of daily cases reported in Pune, Maharashtra and occupation wise infection status of the study population.

## Discussion

Serious setbacks were faced by global healthcare systems due to the COVID-19 pandemic. Coronavirus cases in India increased dramatically in April 2019, reaching a record-breaking daily caseload of more than 40,000.^14^. Indian healthcare workers experienced severe hardships due to the disparity between the number of physicians/ healthcare facilities and the country’s population ^15^. According to the Indian Medical Association (IMA), about 87,000 HCWs contracted the infection, while 573 passed away due to it. Yet another study by the Indian Council of Medical Research (ICMR) suggested that due to staffing shortages in hospitals, 5% of the frontline HCWs, much lower than the present study (∼ 20%), may have developed hospital-acquired COVID-19 infection (HAI) (Murmu 2020).

This study explores the potential associations between various demographic and clinical factors with disease severity and post COVID syndromes among a large cohort of healthcare workers. Asthenia (abnormal physical weakness or lack of energy) emerged as the most common symptom among the long-term COVID symptoms observed. In accordance with a few earlier reports, females were found to have a significantly higher COVID risk than males ^16^,^17^. A few studies have also found a strong correlation between human blood groups and COVID-19 severity and post-COVID symptoms. Blood group B, like previous studies, was found to possess an increased risk of predisposition to long COVID ^17,18^,^19^.

Smokers were discovered to be more vulnerable to COVID-19 infection because of their immunocompromised lung health ^20^). (“The WHO Framework Convention on Tobacco Control (FCTC),” n.d.).^21^. The Global Adult Tobacco Survey 2016-2017 found that 29% of Indian adults (15 years and older) used tobacco, making the country the second-largest consumer of tobacco products, making it a major risk factor for severe COVID (“Tobacco” 2019).

In addition to the above, possible associations of these factors with the severity of the illness were also investigated, but owing to the limiting numbers of severe infections in the study cohort, significant insights were not found.

## Conclusion

Although there exists a handful of primary research exploring potential risk factors associated with COVID-19 infection outcomes and post COVID symptoms, this study is the first of its kind to perform a large-scale cohort analysis of healthcare workers in India. Findings of this study supplements the existing evidence that healthcare workers are at an increased risk of infections, breakthrough infections and reinfections along with the association of increased risk of post COVID syndrome among females, smokers, and individuals with B blood group.

## Data Availability

All data produced in the present work are contained in the manuscript

## Ethics declarations

### Ethics approval and consent to participate

This study was approved by the Institutional Ethics Sub-Committee (IESC) of Dr. D.Y. Patil Medical College Hospital and Research Centre (I.E.S.C/39/2021, Research Protocol No. IESC/FP/2021/33). The participants were explained about the informed consent process as per the approved IESC guidelines.

### Author Contributions

SM helped in the initial ideation of the study. JB, BC, UK, SS, TS, AD, EP, SG, MSS, ZSAS, SB, JV, VYD, JS, AMVK, VK and HSHM were involved in data collection. AVR and MR were involved in data analysis. AG and VS helped in conceptualization of the idea. MR, AG and VS were involved in writing, editing and finalizing the manuscript.

